# Impact of tocilizumab administration on mortality in severe COVID-19

**DOI:** 10.1101/2020.07.30.20114959

**Authors:** Andrew Tsai, Oumou Diawara, Ronald G. Nahass, Luigi Brunetti

## Abstract

**Purpose:** The novel coronavirus disease 2019 (COVID-19) worldwide pandemic has placed a significant burden on hospitals and healthcare providers. The immune response to this disease is thought to lead to a cytokine storm, which contributes to the severity of illness. There is an urgent need to confirm whether the use of tocilizumab provides a benefit in individuals with COVID-19.

**Methods:** A single-center propensity-score matched cohort study, including all consecutive COVID-19 patients, admitted to the medical center who were either discharged from the medical center or expired between March 1, 2020, and May 5, 2020, was performed. Patients were stratified according to the receipt of tocilizumab for cytokine storm and matched to controls using propensity scores. The primary outcome was in-hospital mortality.

**Results:** A total of 132 patients were included in the matched dataset (tocilizumab=66; no tocilizumab=66). Approximately 73% of the patients were male. Hypertension (55%), diabetes mellitus (31%), and chronic pulmonary disease (15%) were the most common comorbidities present. There were 18 deaths (27.3%) in the tocilizumab group and 18 deaths (27.3%) in the no tocilizumab group (odds ratio, 1.0; 95% confidence interval, 0.465 – 2.151; p=1.00). Advanced age, history of myocardial infarction, dementia, chronic pulmonary disease, heart failure, and malignancy were significantly more common in patients who died.

**Conclusion:** The current analysis does not support the use of tocilizumab for the management of cytokine storm in patients with COVID-19. Use of this therapeutic agent should be limited to the context of a clinical trial until more evidence is available.

## Introduction

With the rapid progression of the coronavirus diseases 2019 (COVID-19) pandemic, healthcare providers around the globe have been searching for potential treatment options to combat this disease, which can induce a rapidly progressive and difficult to treat pneumonia that leads to acute respiratory distress syndrome (ARDS). It is caused by a novel coronavirus, severe acute respiratory syndrome coronavirus 2 (SARS-CoV 2). Other highly pathogenic coronaviruses, such as severe acute respiratory syndrome CoV (SARS-CoV) and Middle East respiratory syndrome CoV (MERS-CoV), similarly cause severe pneumonia and often lead to ARDS. Pulmonary inflammation and excessive lung damage have been correlated to a cytokine storm, or aberrant systemic release of cytokines including tumor necrosis factor α (TNF-α), interleukin (IL)-1β, IL-2, IL-6, interferon (IFN)-α, IFN-β, IFN-γ, and monocyte chemoattractant protein-1 (MCP-1).^1,2^ The release of these cytokines can be triggered by the interaction between tumor and effector cells. However, it can also manifest from host immune cells. Clinically, patients with cytokine release syndrome (CRS) present with high-grade fevers, hypotension, and hypoxia.^3^ As a result, therapies that target cytokines or are immunosuppressive are an attractive option to counteract the hyperinflammation.^4^ This assertion is supported by observational data identifying elevated C-reactive protein (CRP), IL-6, and d-dimer as predictors of mortality in this patient population.^5, 6^ Tocilizumab, an IL-6 monoclonal antibody, is currently under investigation as a treatment option for COVID-19 related cytokine release syndrome.^7^ Despite relatively poor quality evidence, tocilizumab has been used around the world.

Although inhibiting the dysregulated inflammatory response may seem intuitive, the complexities that come with blunting the physiologic process associated with fighting infection must be considered. Inflammatory cytokines play a crucial role in the initial innate immune response against viral infections.^8^ Utilizing immunomodulators early on in the treatment algorithm against SARS-CoV-2 may pose more harm than benefit. Administering these pharmacologic agents too late may be in vain as the inflammatory process may have already contributed to irreparable damage to lung tissue. Accordingly, general management with tocilizumab in the institution targeted severe patients prior to the progression of respiratory failure. The objective of this analysis was to evaluate the clinical outcome of in-hospital mortality in patients with COVID-19 treated with tocilizumab.

## Methods

### Study design and participants

A single-center propensity-score matched cohort study, including all consecutive COVID-19 patients, admitted to the medical center who were either discharged from the medical center or expired between March 1, 2020, and May 5, 2020, was performed. Patients were stratified according to the receipt of tocilizumab. All patients received hydroxychloroquine and azithromycin on presentation to the hospital with symptoms consistent with COVID-19 infection. However, azithromycin was held in select patients with underlying cardiac conditions or patients susceptible to cardiac complications. Demographic, clinical, and laboratory data were extracted from the electronic medical records from each of the patients included in the analysis. The baseline (or first available) laboratory values were collected for each patient. At least one ferritin concentration was required for entry into the cohort study, given the relationship with this parameter and cytokine storm.^5, 9^ Further, serum ferritin levels greater than 300 ug/mL were used to justify the use of tocilizumab at our institution. In addition, patients were required to exhibit severe disease at time of administration. Severe disease was defined as an SpO2 ≤ 94% on room air, requiring supplemental oxygen, or requiring invasive or non-invasive mechanical ventilation. The timing and administration of tocilizumab was dependent on the individual clinician; however, the general approach at the institution was to administer tocilizumab early on before the progression of respiratory failure and in patients who met the aforementioned criteria for disease severity. The study was granted expedited approval by the Institutional Review Board (IRB20-20). The reporting of this study conforms to the Strengthening the Reporting of Observational Studies in Epidemiology (STROBE) statement.^10^

### Procedures

Data of demographics, comorbidities, treatments, laboratory results, and clinical outcomes were extracted from the electronic medical record and entered into an electronic dataset for analysis. At least two independent investigators adjudicated study data before analysis. Multiple imputation was used to handle missing data.

### Outcomes

The primary outcome was defined as all-cause in-hospital death.

### Statistical Analysis

Propensity scores were generated using PS Match in SPSS v26.0 (IBM Corporation). Subsequently, propensity score matching was performed to account for treatment strategy influenced by confounding by indication (the tendency of clinicians to prescribe tocilizumab in patients perceived to have cytokine storm and worsening trajectory). Propensity scores were calculated using a multivariable logistic regression model where tocilizumab was the dependent variable. Covariates included in propensity score included age, sex, body mass index, select baseline laboratory values (lactic acid, ferritin, lactate dehydrogenase (LDH), procalcitonin, serum creatinine), hypertension, and comorbidity score. Variables were selected based on clinical significance related to the disease state as well as those that were utilized by prescribers to guide treatment decisions. Controls were matched 1:1 using the nearest neighbor approach without replacement using a caliper width of 0.2. Standardized mean biases were tested to ensure balance after propensity score matching between groups. Categorical data were analyzed using chi-square or Fisher’s exact test as appropriate. The normality of data was assessed through visual inspection of histograms, and Mann-Whitney U. Continuous data were analyzed using the independent samples t-test or Wilcoxon-rank sum as appropriate. The odds ratio and 95% confidence interval were calculated using logistic regression.

## Results

We identified a total of 84 patients who received tocilizumab and 190 patients who were treated without the use of tocilizumab as part of COVID-19 management in our hospital. After the propensity score matching process, each group included 66 patients. Figure 1 provides a schematic representation of the standardized mean difference between covariates selected for propensity score before and after matching. The matching process yielded well-balanced groups, and no significant differences between any of the covariates were observed. Patient demographics and relevant baseline laboratory values are presented in Table 1. The mean age of the study population was 63 ±16.2 years, and 36 patients (27.3%) were females. The most prevalent comorbidities included hypertension (54.5%), diabetes (31.1%), and chronic pulmonary disease (15.2%). At baseline, markers of inflammation including ferritin, CRP, and LDH, were found to be elevated in almost all patients. The mean ferritin was 1027.4 ±957.5 ng/mL, mean CRP was 10.9 ±6.6 mg/dL, and mean LDH was 361.2 ±144.1 U/L. There was no significant difference on the number of patients on a ventilator at baseline between groups (24.2% versus 18.2%; p-0.405, tocilizumab versus control group, respectively). Hydroxychloroquine and azithromycin were commonly used and a similar proportion of patients in each groups received both therapies.

**Table 1.** Patient demographic and clinical characteristics before and after propensity score matching

|  | Unmatched Patients |  |  | Propensity-Score Matched Patients |  |  |
| --- | --- | --- | --- | --- | --- | --- |
|  | Tocilizumab<br>(n=84) | No Tocilizumab<br>(n=190) | p-value | Tocilizumab<br>(n=66) | No Tocilizumab<br>(n=66) | p-value |
| Age (years, mean ± SD) | 61.68 ± 13.55 | 63.58 ± 17.19 | 0.327 | 62.38 ± 13.49 | 61.35 ± 16.09 | 0.691 |
| Female (n, %) | 21 (25.0) | 85 (44.7) | 0.002 | 20 (30.3) | 16 (24.2) | 0.434 |
| Race (n, %) |  |  |  |  |  |  |
| Hispanic | 29 | 87 | 0.082 | 21 (31.8) | 30 (45.5) | 0.108 |
| Caucasian | 23 | 25 | 0.004 | 17 (25.8) | 10 (15.2) | 0.131 |
| African American | 22 | 67 | 0.139 | 20 (30.3) | 21 (31.8) | 0.851 |
| LDH (U/L, mean ± SD) | 429.05 ± 154.50 | 331.19 ± 128.75 | <0.001 | 406.74 ± 143.30 | 379.35 ± 158.93 | 0.300 |
| Lactic acid (mg/dL, mean ± SD) | 1.90 ± 0.93 | 1.74 ± 0.92 | 0.180 | 1.83 ± 0.92 | 1.81 ± 0.98 | 0.863 |
| C-reactive protein (mg/dL, mean ± SD) | 13.01 ± 7.79 | 9.95 ± 5.71 | 0.002 | 11.62 ± 6.61 | 11.83 ± 6.38 | 0.857 |
| Procalcitonin (ng/mL, mean ± SD) | 1.63 ± 7.79 | 0.48 ± 1.45 | 0.177 | 1.14 ± 6.51 | 0.53 ± 1.17 | 0.452 |
| Ferritin (ng/mL, mean ± SD) | 1432.30 ± 1307.96 | 848.35 ± 684.84 | <0.001 | 1146.12 ± 1056.00 | 1249.73 ± 870.27 | 0.540 |
| Serum creatinine (mg/dL, mean ± SD) | 1.30 ± 1.85 | 1.12 ± 1.03 | 0.302 | 1.32 ± 2.06 | 1.25 ± 1.38 | 0.835 |
| Oxygen saturation at baseline (% ± SD) | 86.93 ± 10.74 | 92.04 ± 6.47 | <0.001 | 88.97 ± 8.93 | 89.64 ± 8.12 | 0.654 |
| Body mass index (kg/m<sup>2</sup>, mean ± SD) | 30.47 ± 6.85 | 29.73 ± 7.07 | 0.420 | 30.47 ± 6.72 | 30.00 ± 6.23 | 0.685 |
| Comorbidity index score (mean ± SD) | 1.42 ± 1.62 | 1.18 ± 1.96 | 0.342 | 1.29 ± 1.61 | 1.56 ± 2.57 | 0.466 |
| Hypertension (n, %) | 47 (56.0) | 97 (51.1) | 0.454 | 35 (53.0) | 37 (56.1) | 0.727 |
| Myocardial infarction (n, %) | 9 (10.7) | 16 (17.8) | 0.543 | 7 (10.6) | 6 (9.1) | 0.770 |
| Heart failure (n, %) | 2 (2.4) | 13 (6.8) | 0.161 | 2 (3.0) | 5 (7.6) | 0.440 |
| Cerebrovascular disease (n, %) | 9 (10.7) | 11 (5.8) | 0.148 | 7 (10.6) | 5 (7.6) | 0.545 |
| Peripheral vascular disease (n, %) | 3 (3.6) | 9 (4.7) | 0.761 | 2 (3.3) | 6 (9.1) | 0.274 |
| Dementia (n, %) | 6 (7.1) | 18 (9.5) | 0.529 | 6 (9.1) | 2 (3.0) | 0.274 |
| Chronic pulmonary disease (n, %) | 15 (17.9) | 18 (9.5) | 0.049 | 12 (18.2) | 8 (12.1) | 0.332 |
| Rheumatic disease (n, %) | 2 (2.4) | 7 (3.7) | 0.726 | 1 (1.5) | 3 (4.5) | 0.619 |
| Peptic ulcer disease (n, %) | 0 (0) | 6 (3.2) | 0.182 | 0 (0) | 3 (4.5) | 0.244 |
| Liver disease (n, %) | 1 (1.2) | 3 (0.5) | 1.000 | 1 (1.5) | 1 (1.5) | 1.000 |
| Diabetes (n, %) | 33 (39.3) | 51 (26.8) | 0.039 | 22 (33.3) | 19 (27.3) | 0.573 |
| Hemiplegia/Paraplegia (n, %) | 1 (1.2) | 2 (1.1) | 1.000 | 1 (1.5) | 1 (1.5) | 1.000 |
| Renal disease (n, %) | 6 (7.1) | 6 (3.2) | 0.137 | 3 (4.5) | 4 (6.1) | 1.000 |
| Any malignancy (n, %) | 2 (2.4) | 6 (3.2) | 1.000 | 2 (3.0) | 1 (1.5) | 1.000 |
| Metastatic solid tumor (n, %) | 1 (1.2) | 5 (2.6) | 1.000 | 1 (1.5) | 4 (6.1) | 0.365 |
| AIDS/HIV (n, %) | 0 (0) | 1 (0.5) | 0.670 | 0 (0) | 1 (1.5) | 1.000 |
| Medications |  |  |  |  |  |  |
| Hydroxychloroquine | 75 (89.3) | 163 (85.8) | 0.430 | 60 (90.9) | 59 (89.4) | 0.770 |
| Azithromycin | 51 (60.7) | 121 (63.7) | 0.639 | 39 (59.1) | 43 (65.2) | 0.473 |

<sup>a</sup>Ventilator use is before administration of tocilizumab

**Figure 1:**
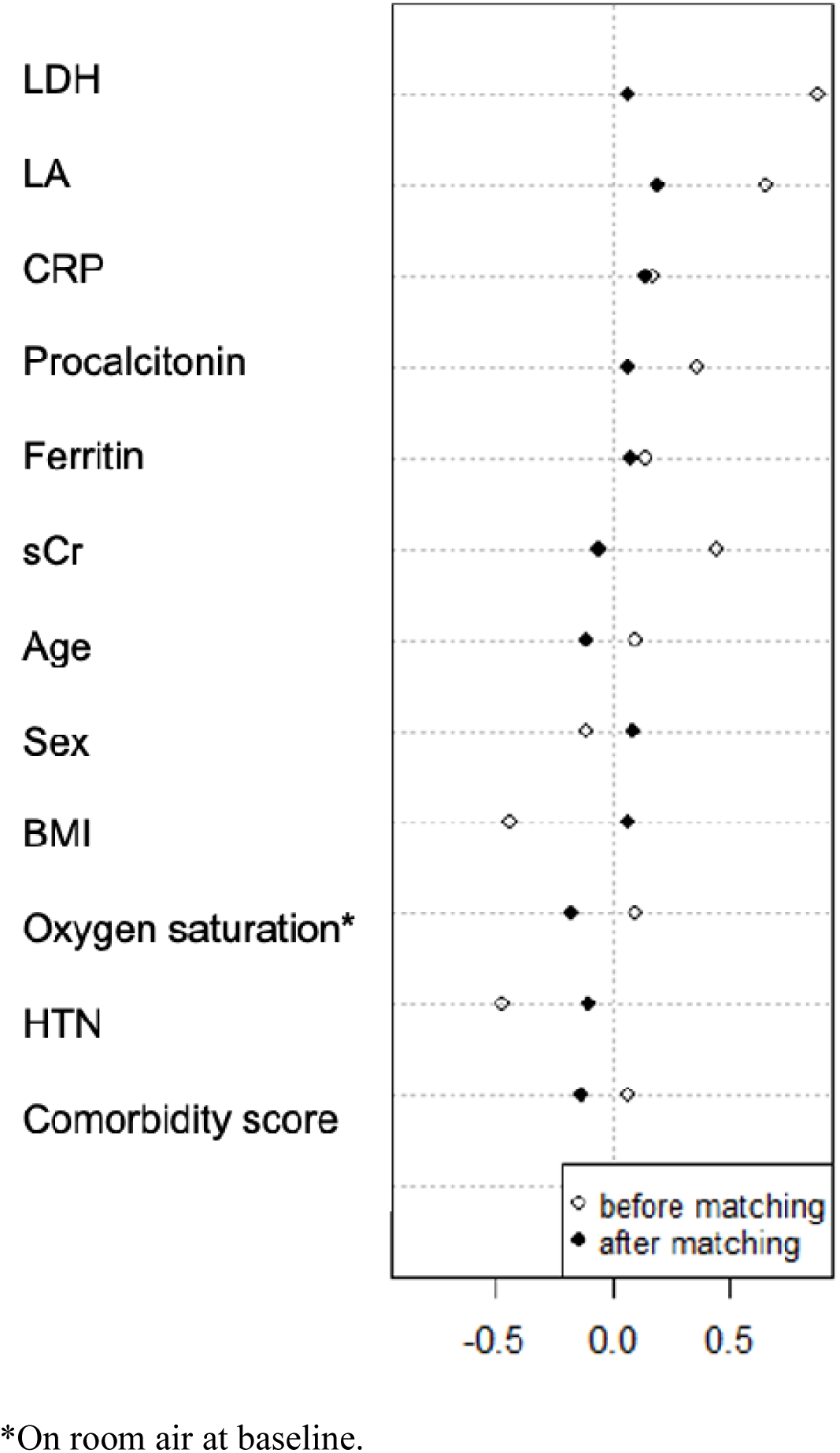
Dot plot displaying the standardized mean difference in covariates included in matching before and after propensity score matching.

Of the patients who received tocilizumab, 10 patients (15.1%) received 800 mg of tocilizumab, 3 patients (4.5%) received 600mg of tocilizumab, and 53 patients (80.3%) received 400 mg. Four patients received a second dose of tocilizumab· In terms of mortality, there were 18 deaths (27.3%) in the tocilizumab group and 18 deaths (27.3%) in the no tocilizumab group (odds ratio, 1.0; 95% confidence interval, 0.465 – 2.151; p=1.00). A secondary analysis using the entire dataset (unmatched), including the propensity score as a predictor in multivariable logistic regression yielded similar results (odds ratio, 0.98; 95% confidence interval, 0.487 – 1.972; p=0.55). In an exploratory analysis of the entire patient population (prior to propensity score matching), characteristics of patients surviving were compared to those who succumbed to the disease process. Table 2 provides a comparison of these covariates between groups. There were significant differences between age, baseline lactic acid, baseline C-reactive protein, baseline oxygen saturation, ventilator use, myocardial infection, heart failure, dementia, chronic pulmonary disease, and any malignancy.

**Table 2.**
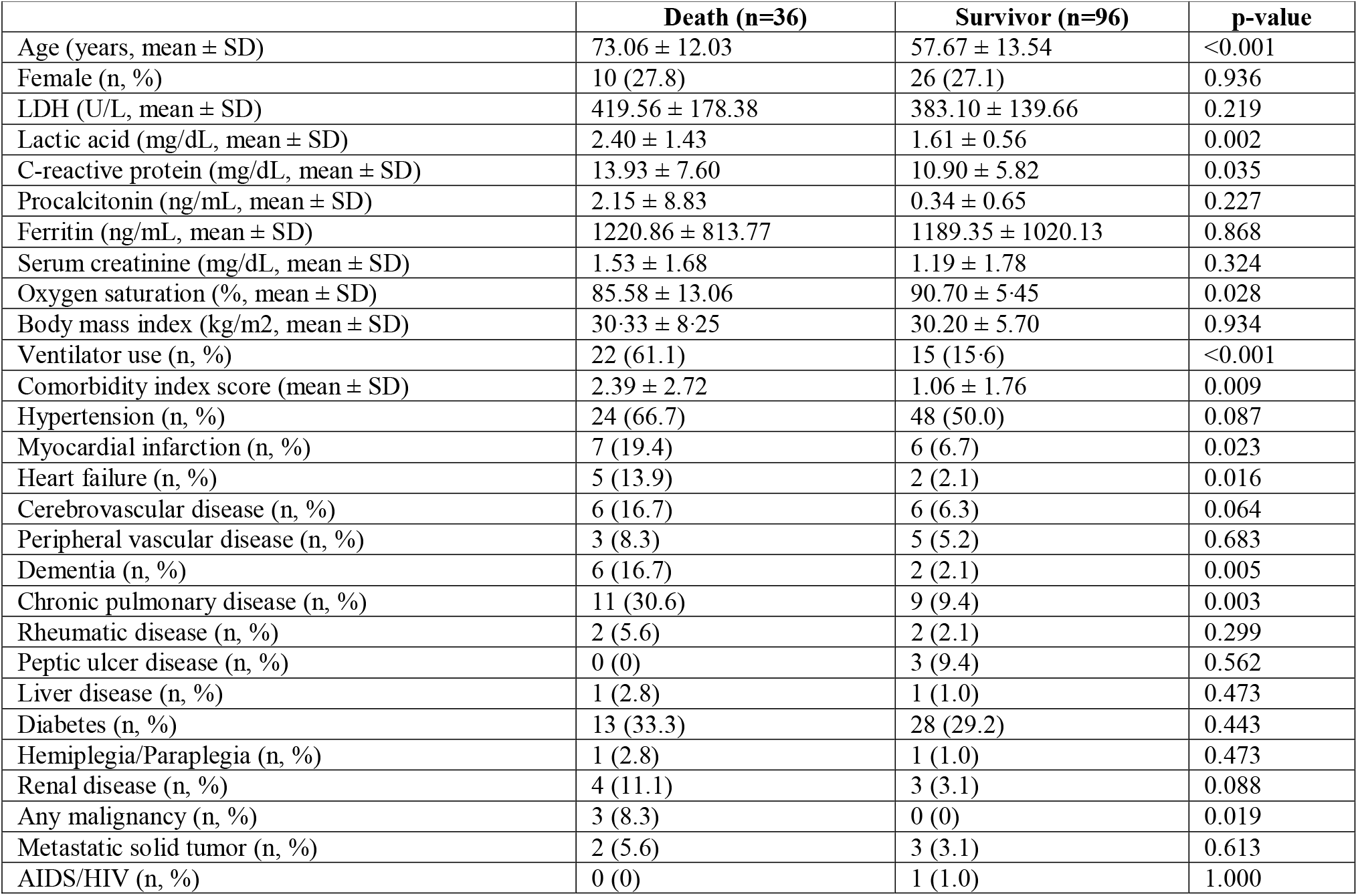
Comparison of patient characteristics between patients surviving and those expiring in the overall population.

## Discussion

Given our current knowledge of the pathophysiology of COVID-19, researchers have proposed repurposing IL-6 receptor antagonists to help curb the cytokine storm. Currently Food and Drug Administration approved for use in the management of rheumatoid conditions and cytokine release storm-related to CAR-T therapy, tocilizumab has gained momentum as a potentially effective option in reducing IL-6 associated fevers and preventing clinical deterioration. Contrary to other observational studies that have been reported, we did not find a benefit with tocilizumab treatment.^11, 12^ There was no difference in mortality in patients treated with tocilizumab versus those receiving supportive care. Although the inflammatory response induced by cytokines may contribute to the severity of illness with SARS-CoV-2 given the essential role that interleukins have in our innate immunity against infectious diseases, our findings suggest that interfering with that response may not be beneficial. Moreover, IL-6 is only one proinflammatory cytokine on a list of many that are released in the downstream pathway of CRS. Important biologic functions of IL-6 include promoting differentiation of B cells into IgM and IgG and stimulating the development and function of Th17 cells.^13^ Specifically, in COVID-19, there is not enough evidence to conclude that inhibiting the signaling of IL-6 alone is enough to quell the ensuing cytokine storm.

We identified several demographic and clinical characteristics that were more common in patients with COVID-19, not surviving. Consistent with other studies, non-survivors in our hospital were older and had more underlying comorbidities, especially cardiovascular and respiratory diseases.^14, 15^ Elderly patients undergo age-related changes that affect their ability to mount an appropriate immune response to emerging infections. Production of new naïve T cells drops significantly between the ages of 40 to 50, threatening the ability of older adults to fight off new viruses like SARS-CoV- 2.16 As our population advances in age, it is also much more common to see individuals with two or more comorbidities. While cardiac disease, bone disease, and malignancy have been determined to increase the risk for poor clinical outcomes in SARS-CoV infections, further studies are needed to explore the mechanisms behind the associations found particularly in COVID-19.^15^

Small observational studies have investigated IL-6 blockade; however, focus on clinical outcomes has been scant.^17, 18^ These studies focused on improvements in laboratory data (i.e., reductions in IL-6, CRP, etc…) with the presumption that improvement in these values leads to clinical benefit. An important consideration in interpreting emerging data is to remain cautiously optimistic.^19^ Surrogate endpoints, while essential to establish or support hypotheses, do not always translate to clinical endpoints. IL-1 is another proinflammatory cytokine that has been linked to increased survival in mice infected with the influenza virus. In addition to suppressing viral replication, IL-1 is involved in the priming of adaptive T and B cells during infection response.^20, 21^ Both early and late phases of virus immunity are influenced by these proinflammatory cytokines. Therefore, extreme caution should be exercised with immunomodulatory therapies that blunt our natural protective mechanisms against disease. These are critical concerns that need to be reiterated as healthcare providers seek therapeutic strategies without strong clinical evidence. While there may be a place in therapy for tocilizumab, it should not be given in the routine management of COVID-19 until more studies are conducted.

There are several limitations to this study. Patients primarily received a single flat 400 mg dose of tocilizumab intravenously, with some receiving a second dose if an additional benefit was considered likely by the treating clinician. The current practice utilizes weight-based dosing for T-cell-induced cytokine release syndrome, and elevations in CRP have been noted to be inversely related to tocilizumab clearance from the body. Therefore, our patients may have been at risk of being underdosed. However, the fixed dosing strategy was based mainly on positive results recently published by Xu and colleagues.^12^ Additionally, serum CRP concentration was included in the propensity score matching. Furthermore, population pharmacokinetic analysis of tocilizumab supports a flat dosing approach due to reductions in the variability of drug exposure among patient weight categories.^22^ Other limitations of our study include the retrospective design, as it may have increased the risk of selection bias toward more severe patients in the tocilizumab arm. While propensity score matching was utilized and resulted in well-matched groups, residual confounding cannot be excluded. Finally, given the small sample size, a type II error is possible. Remarkably, the mortality was identical in both treatment and control groups, suggesting that even if there is a difference in mortality, it may be modest at best.

While the results of the current study were unexpected, recent preliminary reports of randomized controlled trials have dampened enthusiasm for IL-6 antagonizing agents. A clinical trial evaluating sarilumab in COVID-19 patients was discontinued after failing to meet its primary and key secondary endpoints, and instead revealed negative trends in clinical status of patients who were not mechanically ventilated as baseline.^23^ Preliminary analysis of a French tocilizumab study suggested benefit as the study reached its primary endpoint (NCT04331808). While this information provides a reason for pursuit, the primary endpoint was a composite of death or required ventilation. Final data analyses are needed to make any conclusive statements of benefits in terms of overall survival. Clinicians also await completion of the industry sponsored COVACTA trial investigating tocilizumab (NCT04320615). Recently, results from the TOCIVID-19 phase 2 trial reported a 30-day mortality rate of 22.4%, which is comparable to the incidence reported in our results. Although the investigators suggest a potential 30-day mortality benefit with tocilizumab use, the lack of a control group prevents proper evaluation of the benefits for this intervention.^24^ In addition, preliminary results from a randomized controlled trial from Italy of early-stage COVID-19 pneumonia casts further doubt on the efficacy of tocilizumab as it was stopped early due to a lack of efficacy identified at an interim analysis.^25^ Our analysis provides a reason for pause and reset. While there are some data that tocilizumab may be beneficial in a subset of patients, the benefits of this agent are nebulous. Until more data from randomized clinical trials are available, the use of IL-6 directed therapies should occur only in the context of a clinical trial as outlined in the National Institutes of Health treatment guidelines.^26^

## Conclusion

Treatment with tocilizumab in patients with severe COVID-19 was not significantly associated with a difference in in-hospital mortality. Due to the observational nature of our study, these observations require further evaluation in clinical trials to determine the effectiveness of this treatment.

## Data Availability

The data that support the findings of this study are available upon request from the corresponding author, LB. The data are not publicly available due to their containing information that could compromise the privacy of research participants.

## Acknowledgments

We acknowledge all the members of the COVID-19 response team at RWJS for their contributions during the response to the pandemic.

